# Medical students’ and clinical teachers’ experiences and perceptions of using AI simulated patients in psychiatry clinical skills training: a qualitative analysis

**DOI:** 10.64898/2026.10.28.26366085

**Authors:** Dina Gamaleldin, Joanne Rodda, Sukhwinder Shergill, Jim Ang, Sarah Hotham, Verity Williams, Rachel Rice, Hanna Mansi, Sharna Bannet, Olubunmi Olure

## Abstract

**Background:** Simulation-based teaching is central to undergraduate psychiatry education, supporting the development of key clinical skills. Recent advances in generative artificial intelligence (AI), and digital avatars, have enabled the development of AI-driven simulated patients (ASP) that may offer flexible, repeatable learning opportunities. However, there is limited evidence regarding the perceptions of medical students regarding the use of such tools in psychiatry education.

**Aims:** To explore medical student’s attitudes and beliefs towards the use of AI simulated patients for psychiatry clinical skills training, to inform future educational development and integration.

**Method:** Medical students were invited to participate through university mailing lists and departmental and student networks. Interested participants self-enrolled by responding to the invitation. Clinical teachers were recruited through the Royal College of Psychiatrists’ South East Division newsletter. All participants gave both written and verbal consent. A total of sixteen medical students participated across two focus groups. Nine clinical teachers participated across three groups. Each session included a 10-minute hands-on interaction with an exemplar AI based simulation tool, which was developed by the research team. The tool was built in May 2025 using OpenAI’s ChatGPT voice feature (Advanced Voice Mode, powered by GPT-4o), Streamlit (v1.45), and LangChain (v0.3.x). Each AI simulated patients’ interaction was followed by a facilitated group discussion. Sessions were audio-recorded and transcribed verbatim for subsequent thematic analysis.

**Results:** Participants identified both potential benefits and limitations of AI simulated patients in psychiatry education. It was seen as useful for early communication skills training, by offering flexible, low-pressure practice. However, concerns were raised about how well the avatars functioned technically and how accurately they could present people experiencing mental illness, as well as their limited realism compared with actor-based teaching and real patient encounters.

Key priorities for future development included improving emotional realism, expanding scenario options, and ensuring closer alignment with communication skills teaching and institutional systems to support meaningful curriculum integration.

**Conclusions:** Participants recognised the potential of AI simulated patients for undergraduate psychiatry education but identified key barriers to engagement, particularly system reliability and trust, realism representation and institutional access. Addressing these considerations will be important for effective integration, supported by ongoing student involvement.

## Introduction

Student–patient interaction is a principal component of psychiatry education, supporting the development of communication, interviewing, and rapport-building skills (McGaghie) ^1^. However, access to suitable clinical encounters can be limited by placement constraints and restrictions on student involvement in complex or sensitive cases. Actor-based simulation can support the development of clinical skills by offering realistic encounters and structured feedback ^5,6^ and providing a safe environment for practising skills, in addition to exposure to clinical scenarios that may not be consistently encountered in clinical placements ^7^.

Actor based simulation is, however, resource intensive, difficult to scale, and limited to timetabled sessions, restricting opportunities for repeated or independent practice ^8^. Over recent years, digital approaches have emerged, including AI simulated patients, which provide adaptive, naturalistic verbal interaction and represent a shift from earlier, lower-fidelity virtual patient systems (Rodda; Younis) ^9,10^. AI-based virtual patients may now be capable of supporting the development of communication skills, confidence, and diagnostic reasoning (Kononowicz) ^11^. The use of such technologies to support learning aligns with guidance related to technology-enhanced simulation within medical curricula ^12, 13^. Despite this promise, the educational value and acceptability of AI simulated patients in undergraduate psychiatry teaching remains uncertain, with limited research exploring student perspectives and most studies focusing on technological development or other specialties ^14^. As pressures on clinical placements increase due to rising student numbers ^15^, understanding learner attitudes is essential to ensure that any future use of AI simulated patients is responsible, educationally meaningful, and aligned with students’ needs ^16^. This study therefore aimed to explore undergraduate medical student and clinical teacher perceptions, attitudes, and beliefs regarding the use of AI simulated patients in undergraduate psychiatry clinical skills training.

## Participants

Student participants were recruited from Kent and Medway Medical School through newsletters, teaching announcements, and medical school social media. Eligible students were currently enrolled in years 3 or 4 of medical school and were recruited due to their recent exposure to psychiatry simulation teaching.

Clinical teacher participants were recruited through the newsletter of the South East Division of the Royal College of Psychiatrists. Eligible clinical teacher participants were clinicians currently working in psychiatry with experience of undergraduate clinical skills teaching.

Participants gave written informed consent before taking part. Verbal consent was then obtained at the start of every focus group, in two steps. Before any recording began, the facilitator asked the group whether everyone consented to take part, and participants confirmed their agreement verbally, by nodding, or both. Once the audio recording had started, the facilitator asked the group the same question again, so that consent was captured on the recording and appears in the verbatim transcripts. All transcripts were anonymised. No participant declined.

## Data collection

Student focus groups were held both online using Microsoft Teams and in person. Sessions were led by DG, a PhD researcher, with support from an NHS psychiatry resident doctor. Clinical teacher focus groups were conducted online. At the start of each group, all participants spent around 10 minutes using the LLM-ASP, followed by a group discussion guided by the researchers about features, benefits, motivations, barriers, and risks. All sessions were audio recorded, transcribed verbatim, and supported by field notes to capture nonverbal cues and group interactions not evident in the recordings.

## Data analysis

Data were analysed using thematic analysis. Transcripts were coded in NVivo. Codes and developing themes were reviewed through repeated engagement with the data and research team discussions, allowing interpretations to be explored and refined. Initial codes were generated directly from the data and gradually grouped into broader categories and themes. Throughout this process, the research team reviewed the data repeatedly to support clear thinking, reflection, and consistency in interpretation.

The analysis aimed to capture both explicit and underlying patterns within the transcripts, ensuring that the resulting themes remained closely grounded in participants’ own language and experiences. Field notes were used alongside the transcripts to incorporate observations such as tone and affect, allowing for a richer and more contextualised interpretation of participants’ perspectives.

The analytic approach was guided by two complementary frameworks used together. Braun and Clarke’s reflexive thematic analysis ^17^ provided the overarching structure, offering a systematic yet flexible process for generating, reviewing, and refining themes iteratively from the data. Within this broader structure, Saldaña’s multicycle coding approach ^18^ was applied as a practical tool to organise and progressively develop codes across successive rounds of analysis. While Braun and Clarke’s framework directed the wider interpretive process of moving from raw data to meaningful themes, Saldaña’s guidance informed the more granular, line-by-line coding work that underpinned it, which was particularly useful in managing the volume and complexity of data generated across multiple focus groups.

This combined approach is well established in qualitative research, where Saldaña’s coding framework is recognised as a practical complement to Braun and Clarke’s thematic analysis, supporting the systematic organisation of codes before broader interpretive themes are developed ^19^.

## Ethical approval

Ethical approval was granted by the KMMS Research Ethics Committee (ref. GKA200125- 47). The authors assert that all procedures contributing to this work comply with the ethical standards of the relevant national and institutional committees on human experimentation and with the Helsinki Declaration of 1975, as revised in 2013.

## Results

Two student focus groups were conducted; ten students participated in the face-to-face session and six in the online session (table 1), each lasting approximately 60 minutes.

**Table one.** Student Sample Characteristics of Focus Group Participants (n = 16)

| Students | Value |
| --- | --- |
| Total student participants | 16 |
| Focus group format | Face-to-face: 10 participants;<br>Microsoft Teams (online): 6 participants |
| Year of study | Year 3: four students; Year 4: twelve students |
| Education status | All participants were first-degree students |
| Gender | Male: two; Female: 14; Other: 0 |

Nine clinical teachers participated in three 60 minutes online focus groups (table 2).

**Table two.**
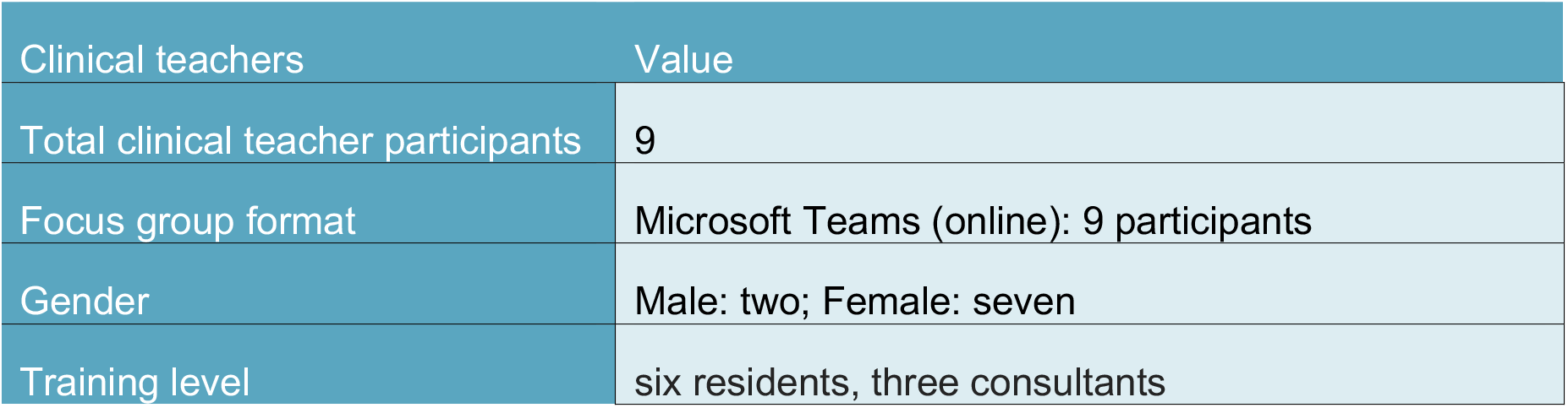
Clinical teachers Participants Sample Characteristics of Focus Group Participants (n = 9)

The thematic analysis for student data identified three overarching themes that captured students’ perceptions and experiences of AI simulated patients in psychiatry education.

### 1. Reflections on the LLM-ASP Interface

Students’ reflections on the system interface centred on issues of trust, realism, and ethical readiness.

A key concern linked to trust was whether these systems could meaningfully represent patients. Participants described limitations in realism and emotional depth, which were viewed as a learning barrier to building rapport and conducting authentic psychiatric interactions:

*‘It’s not real patients, not real conversation*.*’* (P11)

This highlights a gap between AI-generated interactions and real clinical encounters. Students consistently positioned AI simulated patients as a supplementary tool rather than a replacement for existing methods:

*‘AI can never replace actor-based teaching sessions*.*’* (P2)

Other concerns regarding reliability and responsiveness further influenced perceptions of the interface. Students noticed delays and processing issues that disrupted engagement and interaction flow:

*‘It’s got some potential, but maybe it needs a quicker response*.*’* (P4)

Expectations around technological advancement were also evident, particularly the need for more immersive formats:

*‘I just wondered if having a figure, like a video-based AI generated tool not just audio, might be useful*.*’* (P3)

This indicates a preference for visually supported, video-audio interaction rather than audio alone.

Ethical concerns emerged as a central issue within this theme. Students questioned whether these systems could be trusted to handle emotionally sensitive or high-risk content, such as disclosures of self-harm or safeguarding issues, appropriately, given the absence of the professional oversight a real clinician or trained actor would provide:

*‘I would say that ethics could be of concern*.*’ (P6)*

These concerns extended beyond technical functionality to include safeguarding and professional responsibilities. Students questioned what happens to the sensitive content they disclose during practice sessions, including how it is stored, who can access it, and what consent processes govern its use, as well as who holds professional accountability for the AI’s conduct and output within a training context meant to mirror real clinical standards:

‘Like safeguarding and boundaries.’ (P1)

Together, these reflections highlight the importance of ethical, safe, and context-appropriate implementation.

Trust in the system was also linked to institutional validation and integration. Students suggested that credibility could be enhanced through alignment with formal educational structures:

*‘AI needs to be linked to the university portal*.*’* (P5)

### 2. Observations about educational value

Students described the educational value of AI simulated patients in relation to, rather than in replacement of, existing psychiatry teaching methods. Actor-based simulation and real patient encounters remained the preferred and more authentic forms of learning. Students repeatedly reported the limitations of interacting with an artificial system:

*‘It’s not real patients, not real conversation*.*’* (P11)

Students also acknowledged that AI simulated patients could offer a distinct and supplementary form of learning, though concerns emerged about the system’s evidence base being overly broad and insufficiently targeted. Rather than drawing on curated, clinically reliable sources, the LLM was seen as pulling from the internet, with no clear link to vetted or authoritative data:

‘The entire breadth of the internet, not really specific.’ (P14)

Despite these limitations, students recognised potential contribution to preparation and clinical exposure in ways not always achievable through traditional placements. They identified its value in supporting practice for scenarios that may be difficult to access in real clinical settings, particularly where time and resources are limited.

This perceived value was closely linked to opportunities for clinical skill development. Students described how the system could be used to rehearse specific clinical presentations and structured interactions:

*‘I want to practice on certain conditions such as intoxication*.*’* (P7)

This highlights the system’s potential role in targeted skills rehearsal. Students also considered its application to more complex or less frequently encountered cases, including working with specific patient groups in a simulated format. Even in these contexts, however, the lack of realism remained a limiting factor:

*‘Not real patients*.*’* (P10)

An important strength identified by students was the flexibility offered. Students valued the ability to engage with the system independently, outside formal teaching environments, and at their own pace. This was seen as particularly beneficial for revision and self-directed learning:

*‘I want to use it for revision, I’ll use it on my own*.*’* (P8)

### 3. Ethical, Cultural and Governance Considerations

Participants reflected on the broader implications of integrating systems into medical education, particularly in relation to how such technologies may shape future teaching practices and wider professional roles.

Students acknowledged that AI simulated patients could contribute meaningfully to early-stage learning by supporting the development of foundational clinical skills prior to real patient contact:

*‘For early years, it may help guide questioning before patient exposure*.*’* (P4)

This highlights a perceived role for AI tools in scaffolding learning, particularly during the transition from theoretical knowledge to clinical interaction.

Students also expressed concern about the implications of increased reliance on AI-based systems compared to human-led training. Discussions centred on the potential impact on established teaching roles, particularly actor-based simulation and direct patient interaction. Students questioned whether the adoption of AI might reduce opportunities for these forms of learning:

*‘So, no more actors and patient interactions if we start using AI?’* (P9)

The thematic analysis for <u>clinical teachers</u>’ data identified three overarching themes that captured their perceptions and experiences of AI simulated patients in psychiatry teaching.

### 1. Educational Role and Pedagogical Value of AI in Psychiatry Training

Clinical teachers consistently described AI simulated patients as a supplementary learning tool that supports rather than replaces traditional teaching, positioning it as an aid to clinical reasoning and skill development while preserving the learner’s responsibility for leading interactions and decision-making:

*‘It can complement your reasoning, but you still have to lead the whole process*.*’* (P4)

A central advantage identified was the flexibility offering independent, self-paced learning. Clinical teachers explained that students can engage with the tool in their own time and in a more relaxed setting, which may improve confidence when transitioning to real clinical encounters:

*‘Practise now in your own time, more relaxed, to feel more confident, in a real scenario*.*’*

(P2)

Such flexibility supports repeated practice and ongoing skills consolidation outside structured teaching environments.

AI simulated patients was also valued as a safe and supportive environment for early skills development. Clinical teachers highlighted its potential as an introductory step, allowing learners to engage with challenging psychiatric scenarios without the pressure associated with real patient interactions:

*‘This tool could be a first step in a safe teaching environment*.*’* (P4)

This is particularly relevant in psychiatry, where certain clinical presentations may be distressing or unfamiliar for students. One clinical teacher noted that encountering severely unwell patients can be challenging for students and that exposure through AI-based simulation could help:

*‘Prepare them, to understand how it would feel in real life*.*’* (P8)

The role of AI simulated patients was also discussed in relation to curriculum design and assessment. Clinical teachers suggested that its use is most appropriate at later stages of undergraduate training, when students have developed foundational knowledge and are preparing to refine clinical skills.

### 2. Design, Implementation, and Usability of AI simulated patients

Clinical teachers focused on how systems should be designed and implemented to ensure meaningful educational use. A key concern was the current limitation in clinical authenticity, with interactions often perceived as lacking the complexity and natural flow of real psychiatric encounters:

*‘AI is still robotic, it doesn’t give the real feel yet*.*’ (P6)*

Another clinical teacher observed that interactions resembled structured questioning rather than dynamic clinical dialogue:

*‘It seems like it’s kind of interview questions, while in real life it wouldn’t be like this*.*’ (P9)*

This suggests that current systems may not fully capture the relational and adaptive nature of psychiatric practice, where communication depends on spontaneity, emotional interactions, and responsiveness rather than sequential questioning.

Clinical teachers also emphasised that the educational value, depending heavily on the quality of scenario design and ongoing oversight. Well-constructed cases were seen as capable of supporting specific learning objectives. However, challenges could arise when scenarios were not appropriately matched to the learner’s level.

Usability and its impact on communication style was a further concern raised by clinical teachers. The way students interacted was seen as potentially shaping their broader communication behaviours, and in some cases, engagement with the system appeared to encourage more mechanical patterns of interaction:

*‘It almost blunted my communication skills; I became robotic as well*.*’ (P5)*

### 3. Ethical, Cultural, and Governance Considerations

Clinical teachers raised ethical, cultural, and governance considerations that they felt must be addressed to ensure the safe and equitable integration into psychiatric education. A key concern related to equity and representation, particularly the need for systems to reflect the diversity of real patient populations.

Clinical teachers highlighted the importance of including variation in age, ethnicity, and clinical presentation to avoid narrow or stereotypical outputs. Concerns were also raised about default biases embedded within AI systems:

*‘The first thing that the [AI system] is going to create is a man, and a white man’* (P2)

These reflections underlined the participants’ perceived risk of reinforcing existing biases if diversity is not actively and deliberately designed into the system from the outset.

Ethical and responsible use was also a central concern, particularly in relation to how students engage with AI-supported learning. Clinical teachers warned that over-reliance on AI simulated patients could reduce critical thinking and undermine active skill development:

*‘You could be a bad historian and still get everything from the avatar*.*’* (P5)

*‘You rely too much on what something can do for you and so you don’t have to take the effort to learn*.*’* (P2)

These clinical staff views highlight the need for clear pedagogical guidance to ensure that AI tools support rather than replace the development of essential clinical competencies. Transparency about the capabilities and limitations was therefore seen as essential in promoting appropriate expectations and responsible engagement among students.

Clinical teachers also stressed that governance and accountability must be embedded at an institutional level. Responsibility for oversight was viewed as extending beyond individual users, requiring structured integration within university systems and curricula.

Across both groups, six codes recurred consistently: clinical skills development, safe practice environment, complement to existing simulation methods, flexible self-paced learning, impact on medical education, and ethical equity and representation (Figure 3).

**Figure 1.**
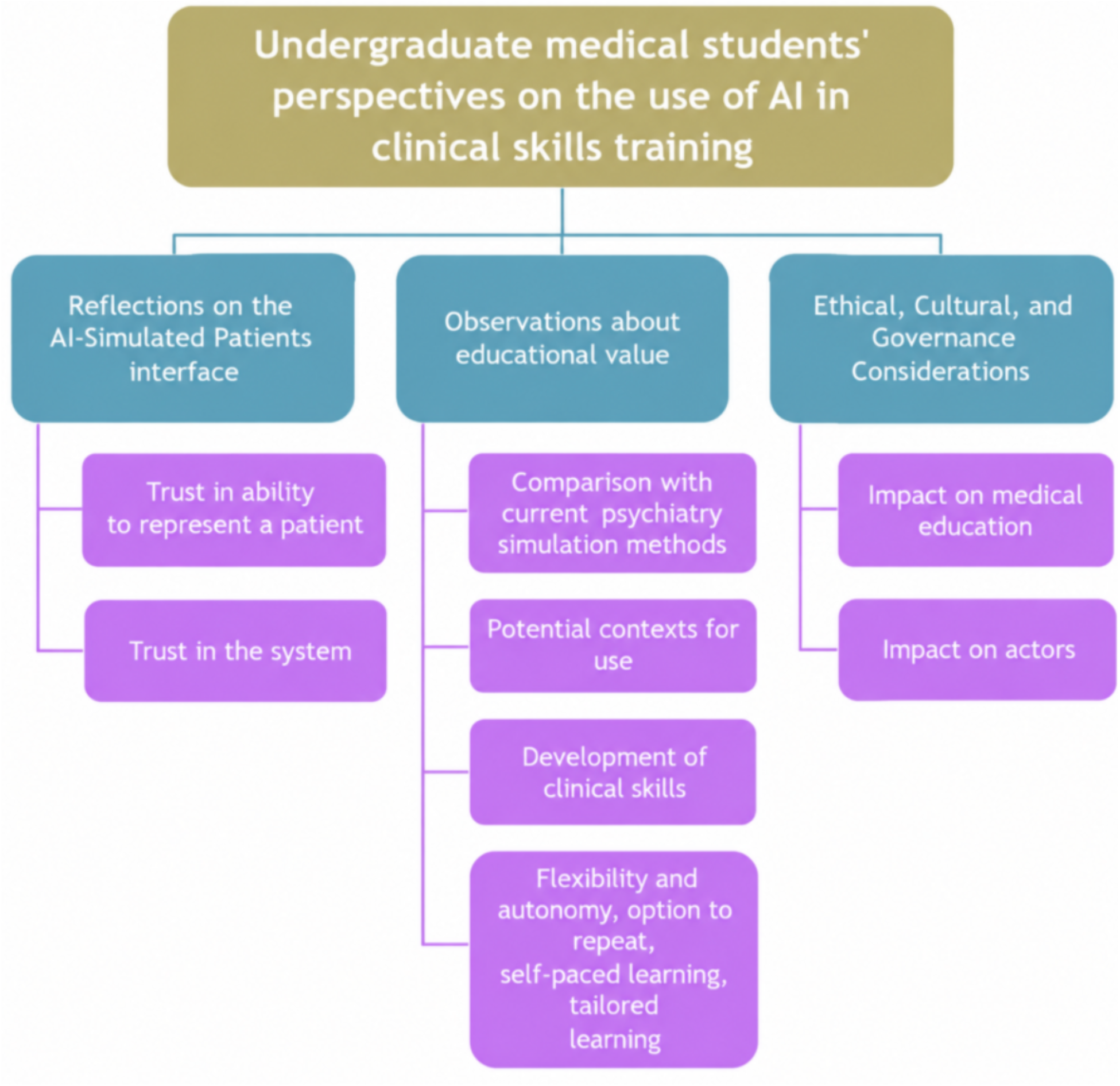
Thematic framework of undergraduate medical students’ perspectives on the use of AI simulated patients in psychiatry clinical skills training: three overarching themes and associated subthemes.

**Figure 2.**
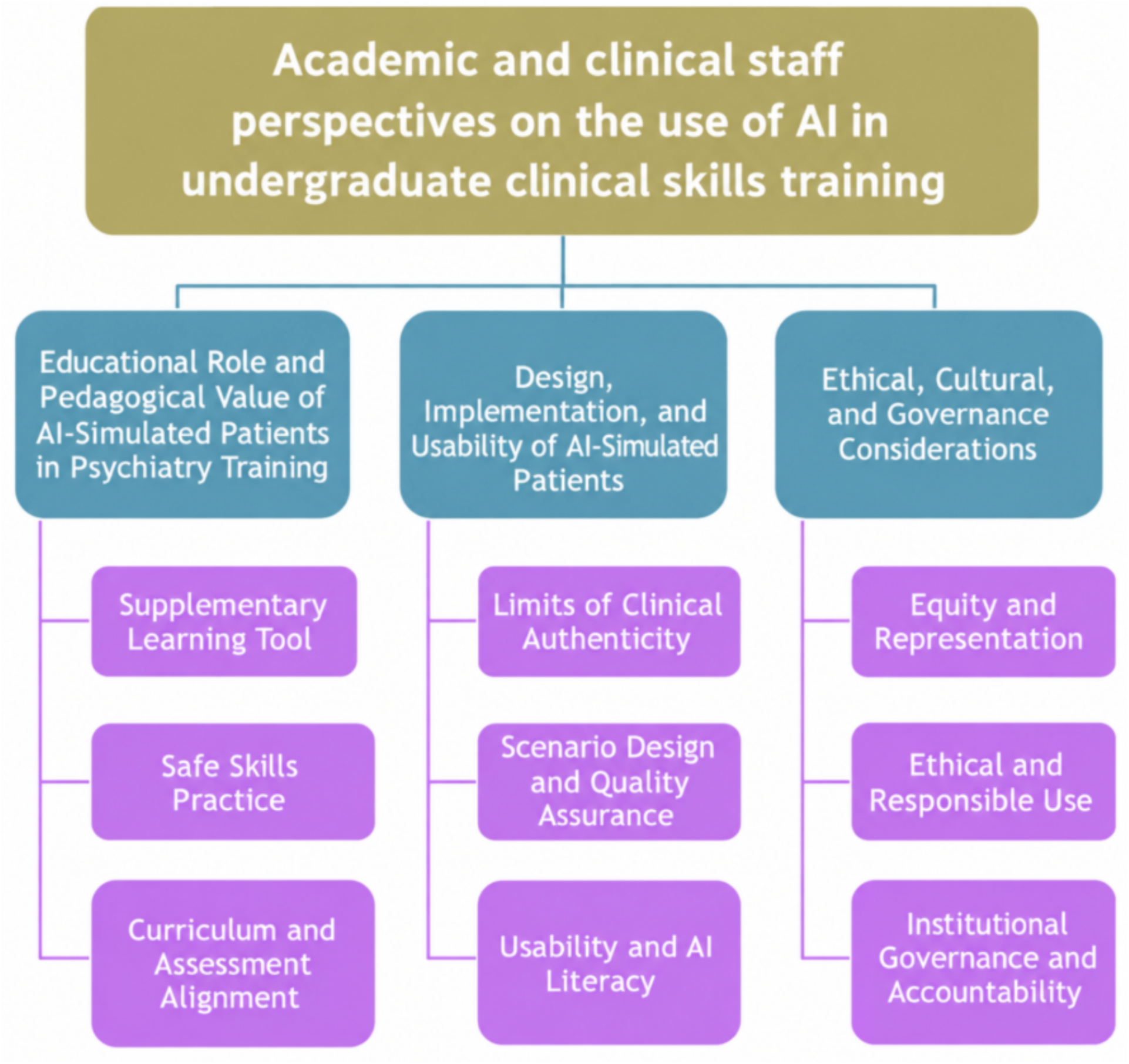
Thematic framework of clinical teachers’ perspectives on the use of AI simulated patients in undergraduate psychiatry clinical skills training: three overarching themes and associated subthemes.

**Figure 3.**
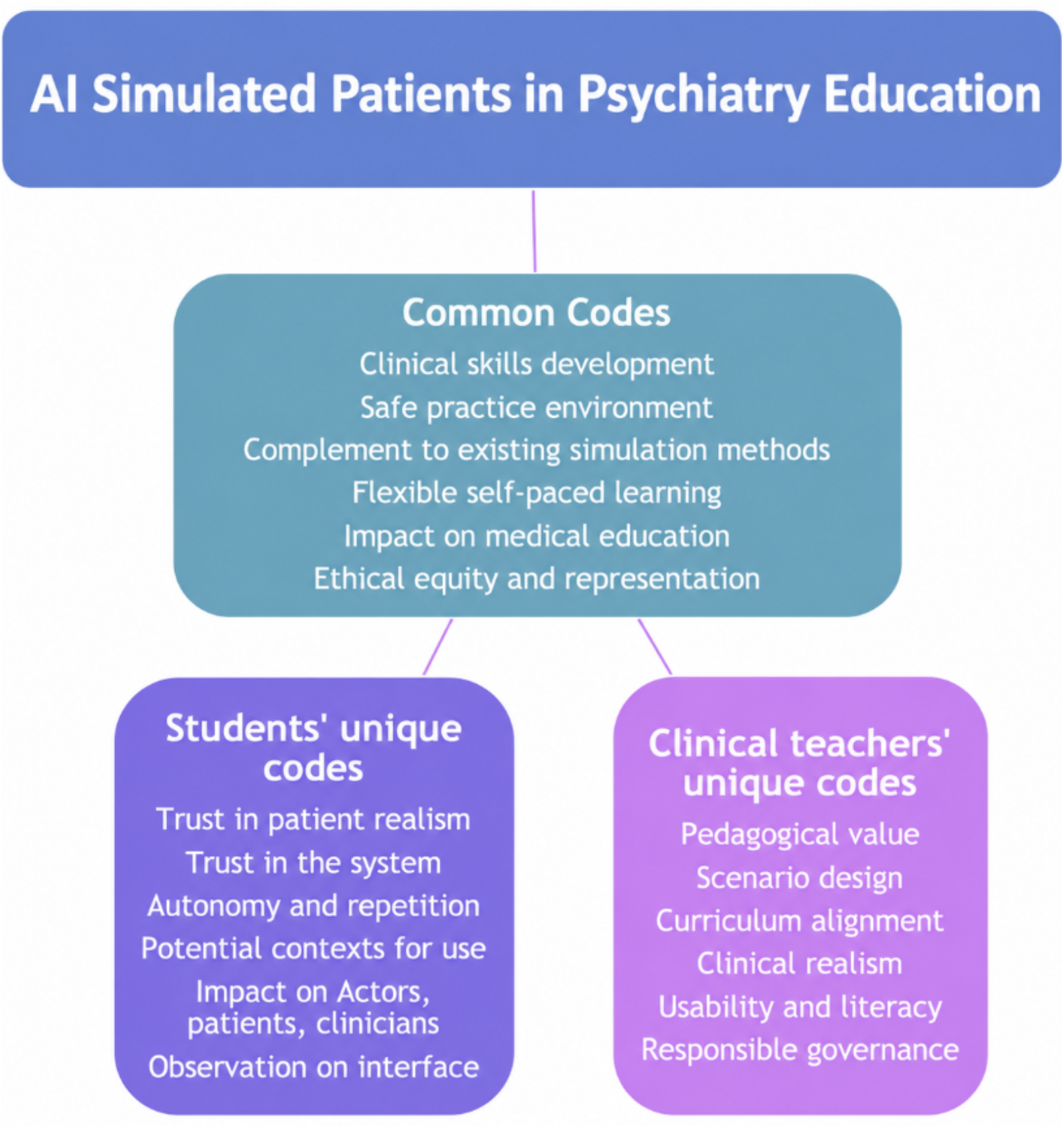
Thematic coding framework for AI-simulated patients in undergraduate psychiatry education. Common codes were identified in both medical student and clinical teacher focus groups, while unique codes emerged from only one group.

Beyond these shared codes, each group also generated themes specific to their own perspective. Students raised additional codes relating to trust in patient realism, trust in the system, autonomy and repetition, potential contexts for use, impact on actors, patients and clinicians, and observations on the interface.

Clinical teachers, in contrast, raised codes concerning pedagogical value, scenario design, curriculum alignment, clinical realism, usability and literacy, and responsible governance. This pattern suggests that while both groups converged on the tool’s core educational value and safety, students focused more on their own experience of trust and usability, whereas clinical teachers focused more on pedagogical integration and oversight.

## Discussion

This study examined perceptions of the use of LLM-ASP among medical students and clinical teachers in undergraduate psychiatry education. Findings show broad agreement that LLM-ASP is a valuable complementary learning tool, though students and clinical teachers differed in emphasis, with students prioritising the learning experience and clinical teachers focusing on evaluation and institutional integration. These findings are discussed in relation to the existing literature below. The findings indicate substantial overlap between students’ and clinical teachers’ perspectives on both the benefits and limitations, alongside differences that reflect their distinct learning and teaching priorities.

This broad agreement suggests that AI simulated patients is perceived as a valuable educational resource across stakeholder groups; however, the observed divergence also implies that its role is shaped by specific pedagogical concerns, with students emphasising the learning experience and educators focusing on evaluation and implementation. This is consistent with Ahsan’s view that the educational value depending on careful implementation and on addressing key challenges to support effective teaching and learning ^20^.

The perspectives of both groups complemented rather than contradicted one another, with each filling gaps left by the other. While students logically framed AI simulated patients primarily as a learning tool, educators’ evaluations reflected broader concerns identified in the literature, particularly around pedagogical value ^21^, curriculum integration ^27^, and ethical governance ^22^. Taken together, these findings indicate that the effectiveness depends not only on the learner and educator experiences, but also on how the tool is integrated within educational and institutional frameworks ^20,21^

AI simulated patients was described by both participant groups as a complementary tool rather than a replacement for traditional medical education, particularly in psychiatry, where emotional sensitivity, relational skills, and empathy depend heavily on human-to-human interaction ^5^. This supports the view that simulation is most effective when combined with direct human interaction, rather than used in isolation, to enhance communication skills and clinical reasoning ^21^. Participants also identified value for independent learning before, during, and after placements, particularly for exam preparation and exposure to rare or complex psychiatric presentations, indicating that the tool may extend wider learning opportunities rather than substitute for clinical experience ^7^.

The overall educational value was recognised by both types of participants. This is in keeping with previous research, particularly that comparing virtual patients with traditional psychiatry teaching methods. Its principal advantages include flexible, self-directed, self-paced learning and opportunities for repeated skills practice, especially in contexts where access to clinical exposure is limited ^23,24^. This highlights the importance of scaffolding scenario complexity in line with student progression and ensuring that cases are designed with a clear pedagogical purpose.

These advantages were recognised within wider medical education and at different stages of medical training, with clinical teachers additionally highlighting the relevance of AI simulated aptients for OSCE and CASC examination preparation. This is in keeping with previous work reporting that AI simulated patients offer meaningful potential for structured, repeatable exam preparation in medical education ^24^.

Psychiatric competency assessments such as these have been noted to permit a degree of variability in examiner judgement, since assessment relies on global judgement rather than fixed checklists ^25^. This variability arguably strengthens the case for tools that allow candidates to build consistent, well-rehearsed practice ahead of these exams.’

Participants also noted the potential value in supporting an increasingly diverse trainee workforce by helping both students and psychiatry residents become more familiar with a range of accents, terminology, and cultural reference points. These shared perceptions align with evidence demonstrating that simulation and deliberate practice enhance skill development, particularly when learners can control the timing and depth of practice ^1^.

Within the common theme of clinical development, both participant groups valued the ability for rehearsing sensitive psychiatric conversations, engaging with a wider range of clinical scenarios, and undertaking preparatory simulation prior to patient contact. These perspectives are consistent with evidence that virtual patients can extend exposure to rare, complex, or less commonly encountered psychiatric presentations ^7^. A related emphasis was placed on the importance of incorporating diverse patient representations, including variations in age, gender, accent, and cultural context. According to Pantziaras this is a widely recognised concept for preparing students for real-world psychiatric practice, suggesting that representational diversity may enhance the authenticity and transferability of learning experiences ^7^.

Considering feedback as a learning support tool, participants highlighted the value of reviewing interaction transcripts alongside receiving immediate, individually tailored feedback aligned with assessment criteria. This aligns with findings by Martins ^26^ who reported that clinical teachers viewed these features as supporting self-assessment, independent learning, and examination preparation related to OSCEs ^26^. These findings reflect established evidence that structured feedback is a vital component of effective simulation-based education and plays a key role in reinforcing skill development ^3^.

System-level improvement through regular testing, evaluation, and structured feedback was identified as a priority by clinical teachers. This aligns with Martin’s emphasis on ongoing development involving both educators and developers to ensure curriculum alignment, maintain clinical accuracy, and support educational relevance and quality assurance with respect to the parallel concept of OSCE developement ^26^. In addition, institutional integration and alignment with formal curricula were viewed by both focus groups as key mechanisms for addressing these concerns, underscoring the need for clinical teacher oversight, validation, and accountability within medical education systems ^27^.

Both groups raised issues relating to secure use, consent, data storage, and information sharing, underscoring the importance of transparent governance and robust data handling in fostering trust and supporting the safe integration into medical education, consistent with existing ethical guidance ^22^.

Usability was another concern raised by participants, particularly students, who emphasised the importance of an intuitive interface and customisable settings for sustained educational engagement.

Trust in AI simulated patients appeared to depend not only on the accuracy of its content but also on how reliably and intuitively the system functioned. This is educationally important, as even relatively minor technical problems may undermine learners’ confidence in the tool and reduce their willingness to use it for repeated, self-directed practice. These findings suggest that usability and technical reliability should be considered alongside clinical and educational accuracy when developing and evaluating AI simulated patients for routine use in medical education ^28^.

Previous research, alongside participant views, has addressed whether Ai simulated patients is ethically ready for medical education, both generally ^29^ and within psychiatry specifically, questioning whether students, educators, and institutions are equipped to handle secure use, consent, data storage, and information sharing appropriately. Students in this present study also expressed uncertainty about the ethical implications and appropriate use of AI in clinical training, questioning both whether the technology was suitable for a high-stakes discipline such as medicine and where the boundaries of appropriate AI conduct should sit, particularly around safeguarding and professional responsibility. These ethical concerns highlight the need for clear governance and robust data management practices to build trust and support the safe adoption in medical education, consistent with existing ethical frameworks ^22^.

Another interesting perspective was students’ comments on the wider effect of the use of AI simulated patients in psychiatry education on actors’ employment, with fear of losing the roles of human actors within the educational system, or having less patient-student interaction, which feeds back to the first common perspective as a complementary, not a replacing, learning tool.

An additional perspective raised by clinical teachers concerned the importance of transparency regarding the capabilities and limitations, who expressed that the clear communication about what these systems can and cannot offer to students is necessary. Understanding system’ s capabilities and limitations is essential to prevent misuse and to manage learners’ expectations, particularly given concerns among clinical teachers that students might develop overreliance on these tools ^30^.

This finding is consistent with calls for medical education to equip learners with the skills required to use and critically evaluate AI within a robust ethical framework ^21^. It also reflects the wider view that AI should augment, rather than replace, human clinical judgement ^32^.

AI-enabled simulation should therefore be integrated into curricula in ways that promote critical evaluation, ethical awareness and appropriate learner autonomy

## Strengths and limitations

This qualitative research explored the views of 16 undergraduate medical students across two focus groups and nine clinical teachers across three focus groups, generating rich accounts of their experiences and perceptions. Iterative coding, repeated engagement with transcripts, and team discussion supported theme development and refinement. Participant diversity in ethnic background and first language was a further strength. However, transferability is limited by the single-site recruitment of students from a well-resourced medical school, the absence of graduate-entry student perspectives, and recruiting all clinical teachers from South-East England.

## Conclusion

Medical students and clinical teachers saw potential value as a flexible and psychologically safe way to practise clinical skills, while consistently viewing it as complementary to, not replacing, actor-based simulation and patient contact. Limited technical reliability and difficulty reproducing the interpersonal and emotional aspects of psychiatric encounters remain important barriers. Participants also raised wider questions around representation and bias, data privacy, safeguarding and professional responsibility, as well as the potential for over-reliance on AI and its impact on existing teaching. These findings suggest that decisions about using AI simulated patients in psychiatry education must consider not only whether the technology works, but how it is introduced, governed and positioned within a curriculum retaining human interaction at its core.

Future research should therefore evaluate both the educational effectiveness and implementation of AI simulated patients within undergraduate psychiatry curricula. Priorities include improving realism and reliability and understanding how these systems can be integrated within university governance and teaching structures. Evaluation should also address ethical issues including data privacy, fairness, bias and appropriate boundaries for AI-supported learning. Given the rapid development of AI technologies, shorter iterative studies may be particularly useful in ensuring that evidence remains relevant to the systems being used.

## Supporting information

COREQ Checklist (Consolidated Criteria for Reporting Qualitative Research)

## Data Availability

All data produced in the present study are available upon reasonable request to the authors

---

| Author | Declaration of Conflict of Interest |
| --- | --- |
| Dina Gamaleldin | No conflicts of interest to declare |
| Prof. Sukhwinder Shergill | No conflicts of interest to declare |
| Dr. Joanne Rodda | No conflicts of interest to declare |
| Prof. Jim Ang | No conflicts of interest to declare |
| Dr. Sarah Hotham | No conflicts of interest to declare |
| Dr. Verity Williams | No conflicts of interest to declare |
| Dr. Rachel Rice | No conflicts of interest to declare |
| Dr. Hanna Mansi | No conflicts of interest to declare |
| Dr. Sharna Bennett | No conflicts of interest to declare |
| Dr. Olubunmi Olure | No conflicts of interest to declare |

