## Supplementary material for "Medical students’ and clinical teachers’ experiences and perceptions of using AI simulated patients in psychiatry clinical skills training: a qualitative analysis": COREQ Checklist (Consolidated Criteria for Reporting Qualitative Research)

**Domain 1: Research team and reflexivity**

| No. | Item | Information reported | Manuscript location |
|---|---|---|---|
| 1 | **Interviewer/facilitator** | Student focus groups were led by DG, supported by an NHS psychiatry resident doctor. | Data collection |
| 2 | **Credentials** | DG holds MA in communication design and a QTS, PGCE in secondary education and is currently undertaking a PhD at Kent and Medway Medical school, The University of Kent. | N/A |
| 3 | **Occupation** | DG was a PhD researcher. The supporting facilitator was an NHS psychiatry resident doctor. | Data collection |
| 4 | **Gender** | DG, Dina Gamaleldin a female researcher | N/A |
| 5 | **Experience and training** | DG received training in qualitative research, including thematic analysis, from Dr Simon Watts. | N/A |
| 6 | **Relationship established before the study** | No prior relationship existed between the researcher and participants. Recruitment was conducted through university networks and the Royal College of Psychiatrists' South East Division newsletter. | N/A |
| 7 | **Participants' knowledge of the researcher** | Participants had no prior knowledge of the researcher before taking part in the study. | N/A |
| 8 | **Researcher characteristics, assumptions and interests** | DG has a background in secondary education and experience facilitating engaging educational sessions. | N/A |

**Domain 2: Study design**

| No. | Item | Information reported | Manuscript location |
|---|---|---|---|
| 9 | **Methodological orientation and theory** | Braun and Clarke's reflexive thematic analysis, supported by Saldaña's multicycle coding approach. | Data analysis |

| | | | |
|---|---|---|---|
| 12 | **Sample size** | 16 medical students across two focus groups and nine clinical teachers across three focus groups. | Results |
| 13 | **Non-participation and withdrawals** | No participants withdrew after enrolment. The number of eligible individuals who declined participation was not recorded. | N/A |
| 14 | **Setting of data collection** | Student groups were held in person and online using Microsoft Teams. Clinical teacher groups were held online. The in-person venue is not specified. | Data collection |
| 15 | **Presence of non-participants** | No individuals other than the participants and research facilitators were present during the focus groups. | N/A |
| 16 | **Description of sample** | Students: four in Year 3 and 12 in Year 4; two male and 14 female; all first-degree students. Clinical teachers: six residents and three consultants; two male and seven female. | Tables 1 and 2 |
| 17 | **Interview or focus-group guide** | Discussion covered features, benefits, motivations, barriers and risks following approximately 10 minutes of interaction with the LLM-ASP. The full guide and whether it was piloted are not reported. | Data collection |
| 18 | **Repeat interviews or focus groups** | No interviews were conducted, no repeat focus groups were conducted. Each participant attended one focus group only. | N/A |
| 19 | **Audio or visual recording** | All sessions were audio-recorded and transcribed verbatim. | Data collection |
| 20 | **Field notes** | Field notes captured non-verbal cues and group interactions and informed interpretation alongside transcripts. | Data collection; Data analysis |
| 21 | **Duration** | Each student and clinical teacher focus group lasted approximately 60 minutes. | Results |
| 22 | **Data saturation** | Data saturation was not assessed, consistent with the reflexive thematic analysis approach. Sample adequacy was considered in relation to the study's focused aims, the relevance of participants' experience and the richness of the focus-group discussions. | N/A |
| 23 | **Transcripts returned to participants** | Transcripts were not returned to participants for comment or correction. | N/A |

**Domain 3: Analysis and findings**

| No. | Item | Information reported | Manuscript location |
|---|---|---|---|

| | | | |
|---|---|---|---|
| **24** | **Number of data coders** | DG coded all transcripts. Developing themes were reviewed and discussed with the research team. | Methods, Data analysis |
| **25** | **Description of coding tree** | Thematic frameworks showing overarching themes and associated subthemes are presented for students and clinical teachers. A detailed coding tree is not described in the text. | Figures 1 and 2 |
| **26** | **Derivation of themes** | Initial codes were generated from the data and progressively grouped into categories and themes through iterative review. | Data analysis |
| **27** | **Software** | NVivo. | Data analysis |
| **28** | **Participant checking of findings** | Participants were not invited to review or provide feedback on the findings during analysis. They will be invited to read the completed paper. | N/A |
| **29** | **Supporting quotations** | Participant quotations are presented with identifiers such as P1 and P2. Student and clinical teacher sections reuse the same identifier format. | Results |
| **30** | **Consistency between data and findings** | Quotations accompany interpretations throughout the Results. Some broader claims would benefit from fuller supporting extracts. | Results |
| **31** | **Clarity of major themes** | Three overarching themes are presented for students and three for clinical teachers. | Results; Figures 1 and 2 |
| **32** | **Clarity of minor themes and diverse cases** | Associated subthemes and differences between student and clinical teacher perspectives are described. Contrasting views within each group are not clearly identified. | Results; Figures 1 and 2 |

Reference: Tong, A., Sainsbury, P. and Craig, J. (2007) 'Consolidated criteria for reporting qualitative research (COREQ): a 32-item checklist for interviews and focus groups', International Journal for Quality in Health Care, 19(6), pp. 349–357. Available at: https://doi.org/10.1093/intqhc/mzm042.
